# Use of heat-not-burn tobacco products, moderate alcohol drinking, and anti-SARS-CoV-2 IgG antibody titers after BNT162b2 vaccination among Japanese healthcare workers

**DOI:** 10.1101/2021.11.29.21267032

**Authors:** Shohei Yamamoto, Akihito Tanaka, Norio Ohmagari, Koushi Yamaguchi, Kazue Ishitsuka, Naho Morisaki, Masayo Kojima, Akihiko Nishikimi, Haruhiko Tokuda, Manami Inoue, Shiori Tanaka, Jun Umezawa, Ryo Okubo, Kunihiro Nishimura, Maki Konishi, Kengo Miyo, Tetsuya Mizoue

## Abstract

**Background:** The effect of heat-not-burn (HNB) tobacco product use and moderate alcohol drinking on immunogenicity to coronavirus disease (COVID-19) vaccines remain elusive. This study aimed to examine the association of tobacco product use and alcohol consumption with anti-SARS-CoV-2 spike IgG antibody titers after the BNT162b2 vaccine.

**Methods:** Participants were 3,457 fully vaccinated healthcare workers in the 4 national centers for advanced medical and research in Japan. Smoking status and alcohol consumption were assessed via a questionnaire, and anti-SARS-CoV-2 spike IgG titers were measured by chemiluminescent enzyme immunoassay using serum collected on the median of 64 days after the second vaccination. Multilevel linear regression models were used to estimate the geometric mean titers (GMT) and the ratios of means (RoM) between groups.

**Results:** Of vaccinated participants, 99.5% (3,440/3,457) were seropositive. Compared with never-smokers (GMT=119), IgG antibody titers were significantly lower among HNB tobacco users (including those who also smoked cigarettes) (GMT=105; RoM=0.88 [95%CI: 0.78–0.99]) and exclusive cigarettes smokers (GMT=96; RoM=0.81 [95%CI: 0.71–0.92]). Compared with non-drinkers of alcohol (GMT=123), alcohol drinkers consuming <1 *go*/day (GMT=114; RoM=0.93 [95%CI: 0.88–0.98]), 1–1.9 *go*/day (GMT=105; RoM=0.85 [95%CI: 0.79–0.93]), and ≥2 *go*/day (GMT=101; RoM=0.82 [95%CI: 0.72–0.94]) had significantly lower antibody titers (*P* for trend<0.01). Spline analysis showed a large reduction of antibody until around 1 *go*/day of alcohol consumption, and then they gradually decreased.

**Conclusions:** Results suggest that in addition to conventional cigarette smoking and heavy alcohol drinking, use of HNB tobacco products and moderate alcohol drinking may be predictors of lower immunological response to COVID-19 vaccine.

**Key Messages:**

- Epidemiological evidence regarding the association of smoking status and alcohol drinking with COVID-19 vaccine-induced antibody levels is scarce.
- Users of heat-not-burn (HNB) tobacco products, as well as cigarettes smokers, had lower antibody titers than never-smokers.
- Not only high-dose but moderate-dose alcohol intake was also associated with decreased vaccine-induced antibody levels.
- HNB tobacco product use and moderate alcohol drinking may be modifiers of COVID-19 vaccine-induced immunogenicity.

## Introduction

With increasing coverage of highly effective mRNA vaccines against severe acute respiratory coronavirus 2 (SARS-CoV-2), the burden of coronavirus disease (COVID-19) is expected to decrease. Studies show a large inter-personal variability in vaccine-induced antibody levels.^1^ While numerous factors may underly the variation, it is crucial to clarify modifiable factors that influence post-vaccine immunogenicity. Smoking and heavy alcohol drinking are known to interfere with the activation of innate and acquired immunity ^2, 3^ and may thus suppress vaccine-induced antibody production.

Epidemiological evidence regarding the association of smoking and alcohol use with post-vaccine antibody titers has been inconsistent.^4-7^ No data are available linking post-vaccine antibody response to the use of heat-not-burn (HNB) tobacco, which has been increasingly popular in global market.^8^ Similar to conventional cigarettes, HNB tobacco products contain nicotine,^9^ which may harm the immune system.^2^ Regarding alcohol drinking, previous studies did not address the dose-response relationship, with attention to the effect of moderate drinking, which has been hypothesized to enhance vaccine response.^10^ This would be a critical issue for East Asians including Japanese, who have a high prevalence of genetic mutation in an alcohol-metabolizing enzyme.^11^

The present study sought to examine the association of smoking (including use of HNB tobacco) and alcohol consumption with IgG antibody titers against SARS-CoV-2 spike protein among the staff of national medical research centers in Japan who received two doses of BNT162b2 vaccine.

## Methods

### Study design

In 2020, a multi-center collaborative study comprising repeat serological surveys was launched among the workers of the six National Centers for Advanced Medical and Research in Japan (6NC). Each NC performs the survey at least once per year during the COVID-19 epidemic. Prior to each survey, the researchers agreed on the survey procedure, including questionnaire and antibody assays used across 6NC, considering the then-current situation of the COVID-19 epidemic, commercial availability of antibody assays, and vaccination program. Written informed consent was obtained from each participant. After completing the opt-out process, the survey data were anonymized and submitted to the study committee for pooled analysis. The study design and procedure for data collection were approved by the ethical committee of each center, while those of pooling study were approved by that of the National Center for Global Health and Medicine (NCGM) (the approved number: NCGM-G-004233).

### Study population

For the current study, we analyzed data from 4 national centers, which performed a serological survey between June and August 2021 after the completion of the vaccination program for the staff and submitted the data for pooling analysis. The details of the survey and vaccination schedule for each national center were described in **Supplemental Table 1**.

Of 8,190 workers invited to the survey, 5,718 participated (70%). Of these, 5,013 reported receiving two doses of the COVID-19 vaccine (BNT162b2, Pfizer-BioNTech) by the survey. Of these, we excluded those who attended the survey for the blood draw within 14 days of the second vaccination (n=193), those who had extremely low antibody titers (<1 SU/mL) (n=5), those who lacked data on smoking status or alcohol consumption (n=1,346), and those who lacked data on covariates (n=12), leaving 3,457 for analysis.

### Data collection

We asked participants to donate venous blood and answer a questionnaire including queries on smoking, alcohol consumption, vaccination, the history of COVID-19, comorbid conditions (hypertension, diabetes, and cancer), leisure-time physical activity, and sleep duration. For smoking conventional cigarettes, we asked participants to select one of the following options: never, quit, smoking less than daily, smoking ≤10, 11–20, or ≥21 cigarettes/day. We also asked the use of new tobacco products (HNB tobacco products [IQOS, glo, PULZE, WEECKE, etc] or e-cigarette) in the last month. Based on the answers to these questions, we categorized the participants into five groups: never smoker (never smoked cigarette and did not use HNB tobacco products), former smoker (quit smoking cigarette and did not use HNB tobacco products), current smoker who used only HNB tobacco products, current smoker who smoked only conventional cigarettes, and current smoker who used both conventional cigarettes and HNB tobacco products (dual users). We did not consider e-cigarette use because most e-cigarettes available in the Japanese market do not contain nicotine. In post hoc analysis, we combined the categories of exclusive HNB tobacco users and dual users in one to increase statistical power.

For alcohol consumption, we asked the frequency of alcohol intake (ranging from never to daily) and the amount of intake per ocassion (ranging from <0.5 to ≥4 *go*/day). In Japan, *go* (180 mL) is used as the conventional unit to measure alcohol amount; 1 *go* of Japanese *sake* contains approximately 23 g of ethanol, which is equivalent to 500 mL of beer, 110 mL of shochu (25% alcohol content), double (60 mL) of whisky, or 180 mL of wine. We calculated averaged daily consumption and classified the participants into five groups: non-drinker, occasional drinker (1–3 days/month), and weekly drinker consuming <1 *go*/day, 1–1.9 *go*/day, or ≥2 *go*/day.

### Serological assay

We quantitatively measured SARS-CoV-2 IgG antibodies against spike protein in serum using the chemiluminescence enzyme immunoassay (CLEIA) platform (HISCL) manufactured by Sysmex Co. (Kobe, Japan). The sensitivity and specificity were reported as 98.3% and 99.6%, respectively.^12^ We confirmed that IgG antibody titers measured on this assay were highly correlated with those measured on the AdviseDx SARS-CoV-2 IgG II assay (Abbott ARCHITECT^®^) using data of two centers that measured antibodies with both assays (n=2,961): spearman’s rho = 0.96 (95% confidence intervals [CI]: 0.95–0.96) (**Supplemental Figure 1**).

### Statistical analysis

We performed multilevel linear regression analysis to examine the associations of smoking and alcohol use with anti-spike IgG antibody titers in log-scale. Fixed-effect covariates included age (continuous), sex, job category (doctor, nurse, allied healthcare professional, administrative staff, researcher, and other), body mass index (BMI) (continuous), an interaction term between sex and BMI,^13^ the interval between the second vaccination and blood sampling (continuous), and a square term of the interval, the history of COVID-19, comorbid conditions (diabetes, hypertension, and cancer), leisure-time physical activity (0, 1 to 59, or ≥60 min/wk), sleep duration (<6, 6 to 6.9, or ≥7 h), smoking status, and alcohol consumption. The random-effects intercept term was set by national center level. The estimated effects of exposures were back-transformed to present the geometric mean titer (GMT) and ratios of means. We tested the statistical significance between groups by comparing the ratios of means. We drew a restricted cubic spline curve to visualize the dose-response association between alcohol consumption and IgG antibody titers, with 3 knots placed at the 5^th^, 50^th^, and 95^th^ percentiles as recommended.^14^ Statistical analysis was performed using Stata 17.0 (StataCorp LLC). All *P* values were 2-sided, and *P* < 0.05 was considered statistically significant.

## Results

The median age of participants was 41 years (interquartile [IQR]: 30–50 years), 72% were women, 0.5% had a history of COVID-19, and the median BMI was 21.4 (IQR: 19.6–23.5). One-third were nurses (34%), followed by allied health professionals (18%), administrative staff (15%), doctors (14%), and researchers (13%). Of 212 current smokers (6.1% of the participants), nearly half (53%) used HNB tobacco products. Thirty nine percent drank alcohol beverage at least once per week. The median interval between the second vaccination and blood sampling was 64 days (IQR, 38–74 days). The median titers of anti-SRAS-CoV-2 spike IgG antibody were 144 SU/mL (IQR: 77–249), with 99.5% (3,440/3,457) of participants were seropositive (threshold ≥10 SU/mL) (**Table 1**).

**Table 1.**
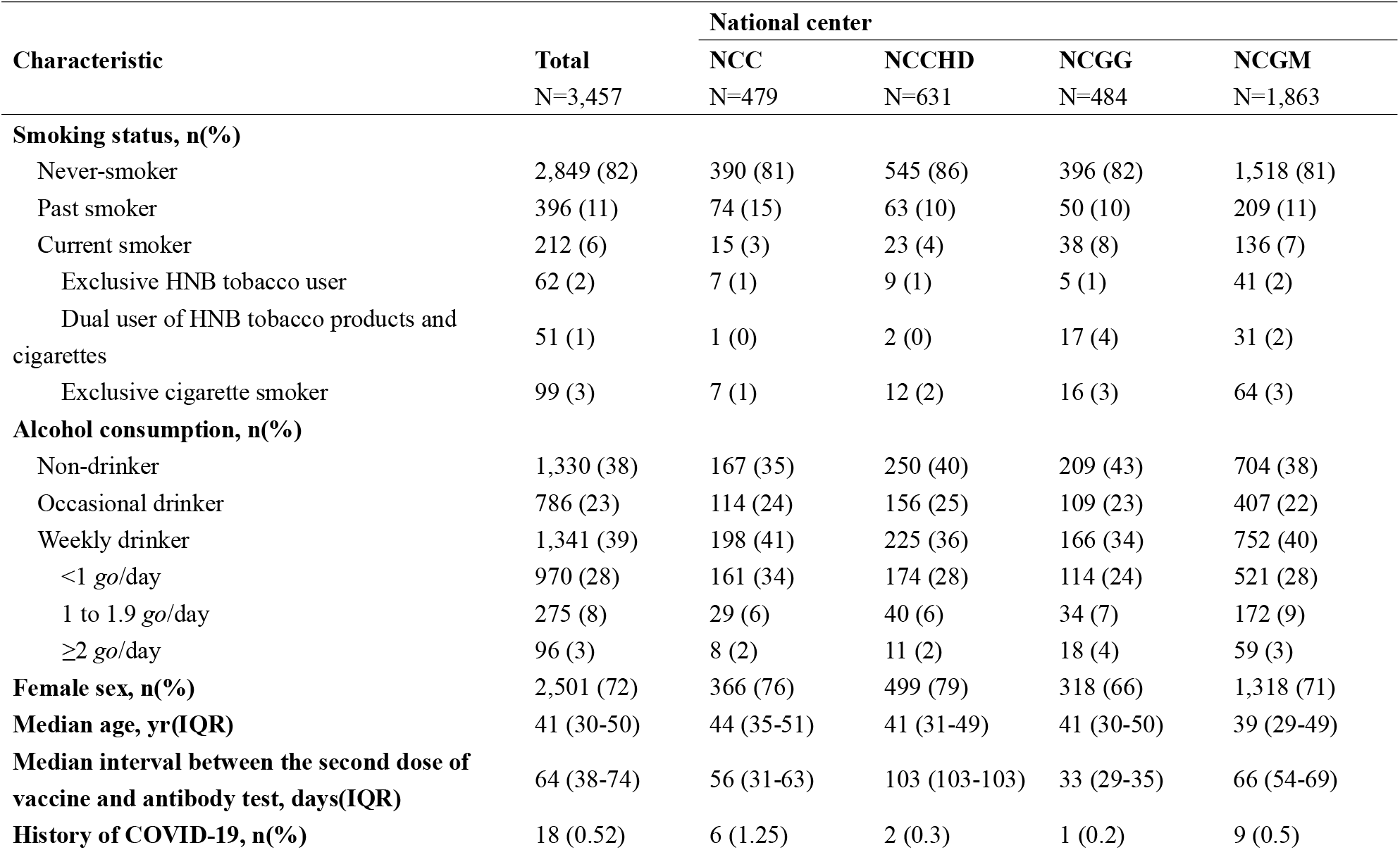
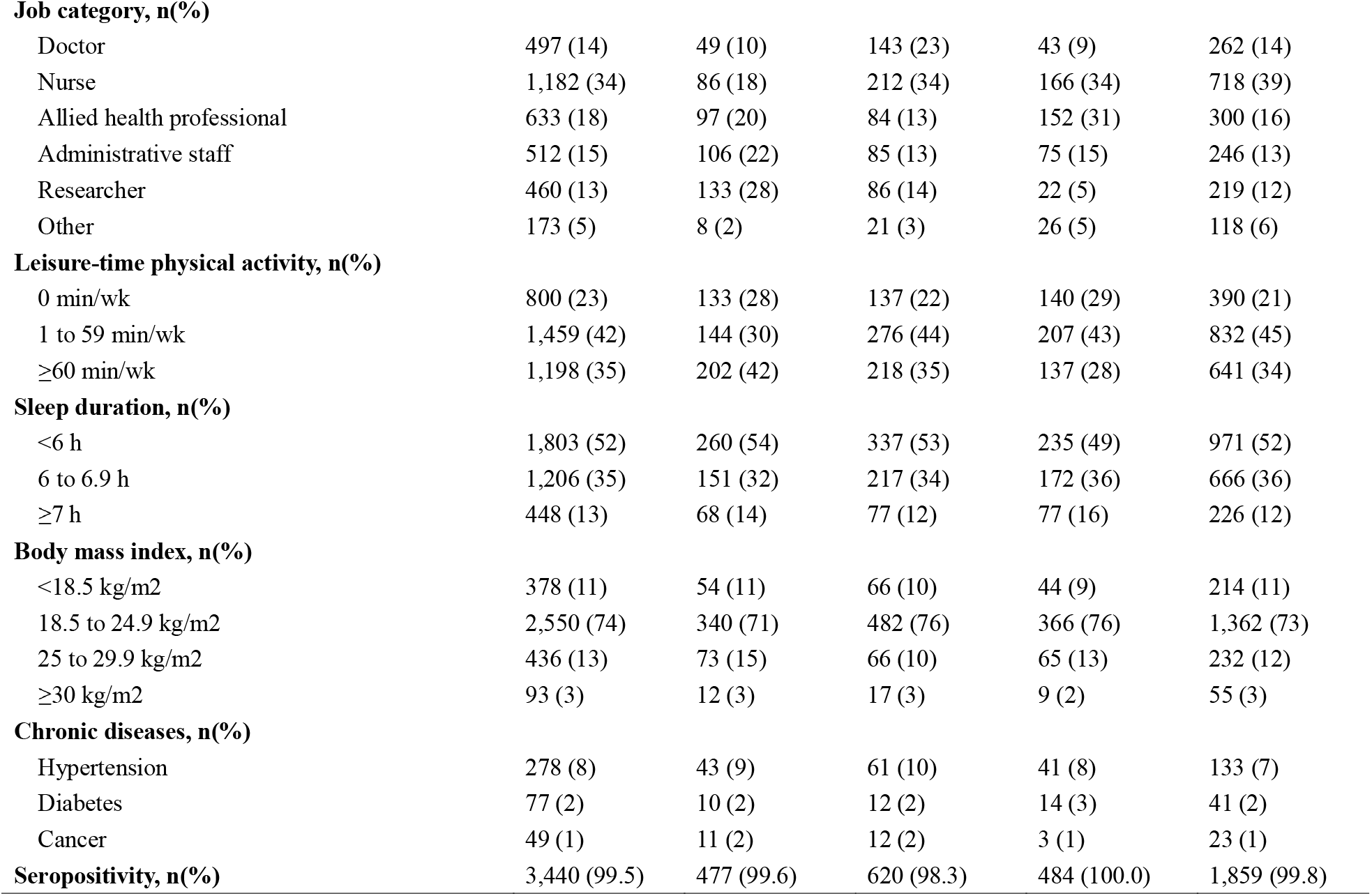
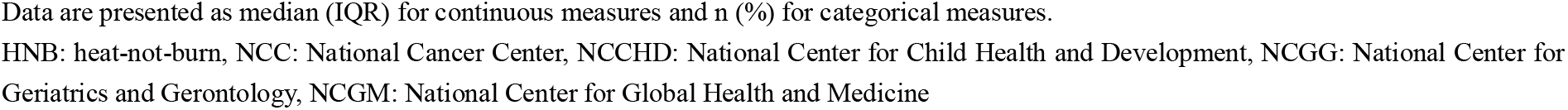
Characteristics of participants according to national center

Current smokers using any tobacco product had lower antibody titers (GMT, 101; ratio of mean, 0.85 [95% CI: 0.77–0.93]) compared with never-smokers **(Supplemental Table 2**). As shown in **Table 2**, exclusive cigarette smokers had significantly lower GMT than never-smokers (GMT, 119 versus 99; ratio of means, 0.81 [95%CI: 0.71–0.92]). Exclusive HNB tobacco product users and dual users also showed similarly lowered GMT (103 and 108, respectively), although the differences from never-smokers were not statistically significant (ratio of means, 0.87 [95%CI: 0.74–1.02] and 0.91 [95%CI: 0.76–1.08], respectively). In the post-hoc analysis combining the two categories of HNB tobacco users (n=113), the reduction reached statistical significance (GMT, 105; ratio of mean, 0.88 [95%CI: 0.78–0.99]). Among daily cigarette smokers, those consuming ≥11 cigarettes per day showed a greater reduction in IgG titers than those consuming <11 cigarettes per day; GMTs (ratio of means) were 92 (0.77 [95%CI: 0.62–0.95]) and 104 (0.87 [95% CI: 0.76–1.00]), respectively **(Supplemental Table 2**).

**Table 2.**
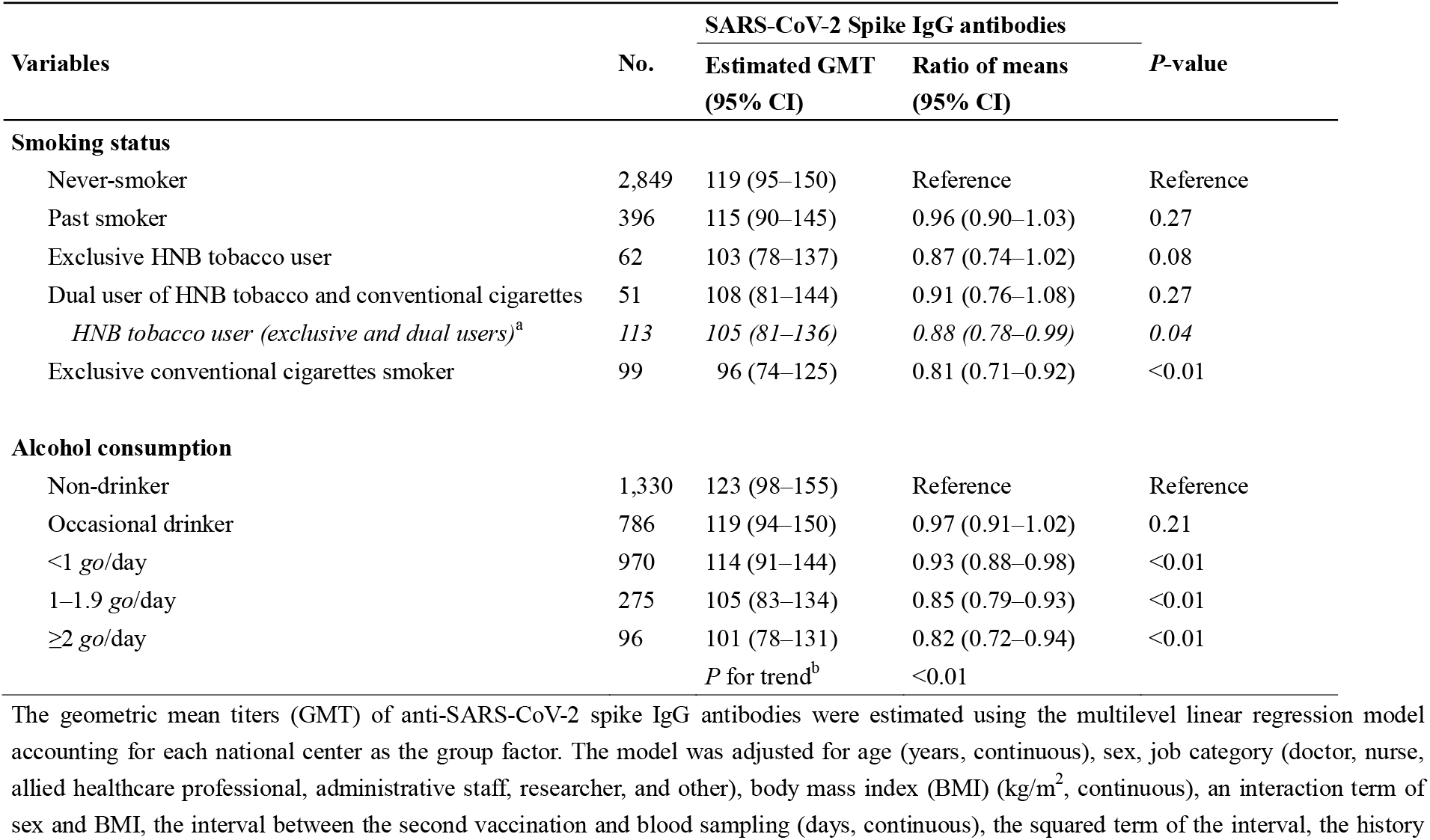
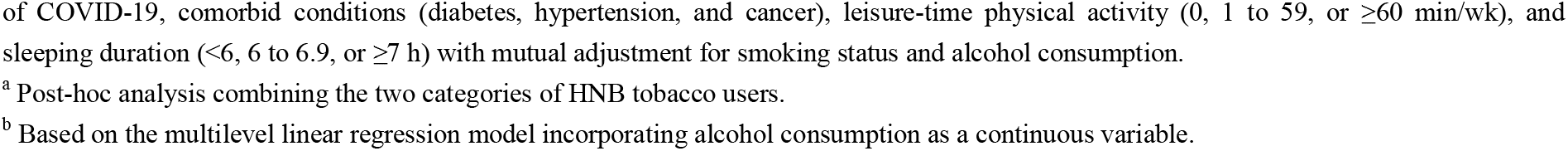
The estimated geometric means (GMT) with 95% confidence intervals (CI) with smoking status and alcohol consumption among 3,457 vaccinated healthcare workers.

| Variables | No. | SARS-CoV-2 Spike IgG antibodies |  | *P*-value |
| --- | --- | --- | --- | --- |
|  |  | Estimated GMT<br>(95% CI) | Ratio of means<br>(95% CI) |  |
| **Smoking status** |  |  |  |  |
| Never-smoker | 2,849 | 119 (95–150) | Reference | Reference |
| Past smoker | 396 | 115 (90–145) | 0.96 (0.90–1.03) | 0.27 |
| Exclusive HNB tobacco user | 62 | 103 (78–137) | 0.87 (0.74–1.02) | 0.08 |
| Dual user of HNB tobacco and conventional cigarettes | 51 | 108 (81–144) | 0.91 (0.76–1.08) | 0.27 |
| *HNB tobacco user (exclusive and dual users)<sup>a</sup>* | *113* | *105 (81–136)* | *0.88 (0.78–0.99)* | *0.04* |
| Exclusive conventional cigarettes smoker | 99 | 96 (74–125) | 0.81 (0.71–0.92) | <0.01 |
| **Alcohol consumption** |  |  |  |  |
| Non-drinker | 1,330 | 123 (98–155) | Reference | Reference |
| Occasional drinker | 786 | 119 (94–150) | 0.97 (0.91–1.02) | 0.21 |
| <1 *go*/day | 970 | 114 (91–144) | 0.93 (0.88–0.98) | <0.01 |
| 1–1.9 *go*/day | 275 | 105 (83–134) | 0.85 (0.79–0.93) | <0.01 |
| ≥2 *go*/day | 96 | 101 (78–131) | 0.82 (0.72–0.94) | <0.01 |
|  |  | *P* for trend<sup>b</sup> | <0.01 |  |

The geometric mean titers (GMT) of anti-SARS-CoV-2 spike IgG antibodies were estimated using the multilevel linear regression model accounting for each national center as the group factor. The model was adjusted for age (years, continuous), sex, job category (doctor, nurse, allied healthcare professional, administrative staff, researcher, and other), body mass index (BMI) (kg/m<sup>2</sup>, continuous), an interaction term of sex and BMI, the interval between the second vaccination and blood sampling (days, continuous), the squared term of the interval, the history
of COVID-19, comorbid conditions (diabetes, hypertension, and cancer), leisure-time physical activity (0, 1 to 59, or $\geq 60$ min/wk), and sleeping duration ( $<6$ , 6 to 6.9, or $\geq 7$ h) with mutual adjustment for smoking status and alcohol consumption.
<sup>a</sup> Post-hoc analysis combining the two categories of HNB tobacco users.
<sup>b</sup> Based on the multilevel linear regression model incorporating alcohol consumption as a continuous variable.

Weekly drinkers of alcohol beverage had significantly lower antibody titers than non-drinkers (**Supplemental Table 2**). Spike IgG antibody titers steadily decreased with increasing amount of alcohol consumption. The estimated GMTs of the antibody for non-drinkers, occasional drinkers, and weekly drinkers consuming <1 *go*/day, 1–1.9 *go*/day, and ≥2 *go*/day were 123 (95%CI: 98–155), 119 (95%CI: 94–150), 114 (95%CI: 91–144), 105 (95%CI: 83–134), and 101 (95%CI: 78–131), respectively (P for trend <0.01) (**Table 2**).

Compared with non-drinkers, antibody titers were statistically significantly lower even among moderate drinkers consuming <1 *go*/day (<23g ethanol), with the ratio of means for 0.93 (95%CI: 0.88–0.98). Spline analysis yielded a clear dose-response relationship, showing a large reduction in the ratio of means until around 1 *go*/day of alcohol consumption, followed by gradual decreasing trend with increasing alcohol consumption (**Figure 1)**.

**Figure 1.**
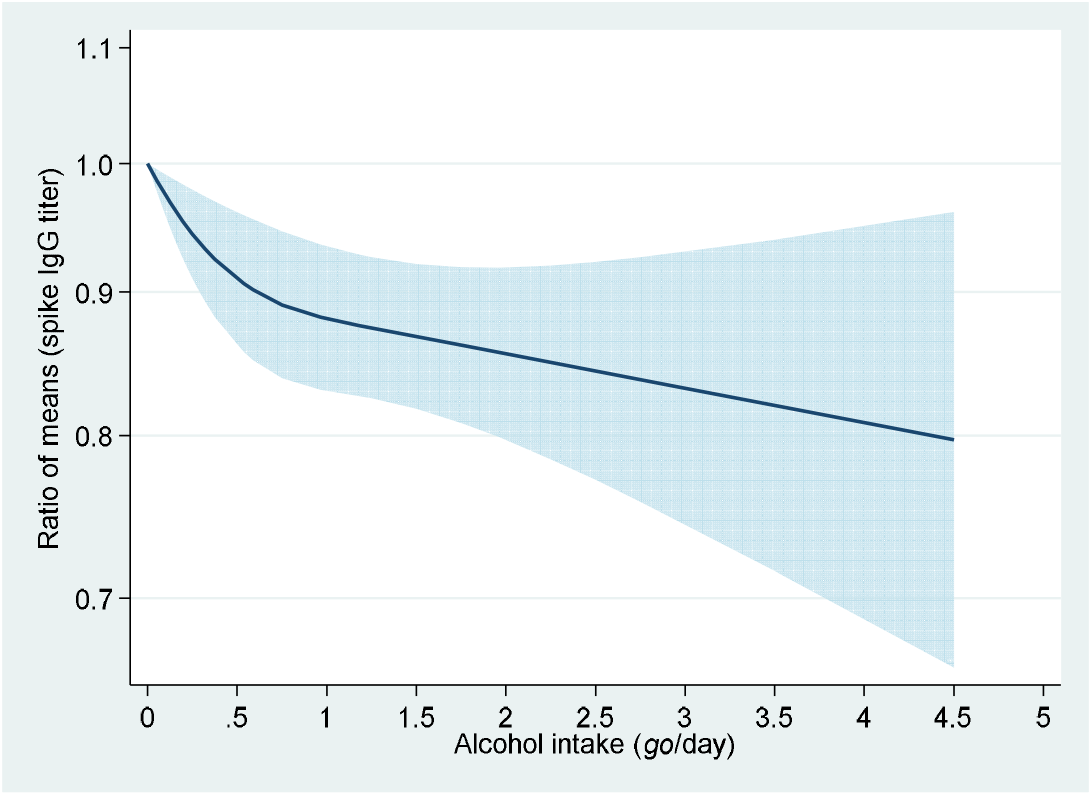
Dose-response association between alcohol consumption and anti-SARS-CoV-2 spike IgG titers The y-axis is the ratio of means with the shaded area representing 95% confidence intervals (linear trend, P<0.01; non-linear trend, P=0.02). The reference value is 0 *go*/day (non-drinkers). The model was adjusted for age (years, continuous), sex, job category (doctor, nurse, allied healthcare professional, administrative staff, researcher, and other), body mass index (BMI) (kg/m^2^, continuous), an interaction term of sex and BMI, the interval between the second vaccination and blood sampling (days, continuous), the squared term of the interval, the history of COVID-19, comorbid conditions (diabetes, hypertension, and cancer), leisure-time physical activity (0, 1 to 59, or ≥60 min/wk), sleeping duration (<6, 6 to 6.9, or ≥7 h), and smoking status. One *go* of Japanese sake contains approximately 23 g of ethanol.

## Discussion

In 3,457 staff of national medical research centers in Japan who completed two-dose of the COVID-19 BNT162b2 vaccine, HNB tobacco products users (exclusive or combined with cigarettes) and exclusive cigarettes smokers had significantly lower anti-SARS-CoV-2 spike IgG antibody titers compared with never-smokers. There was a clear decreasing trend of antibody titers with increasing alcohol consumption, with significant reduction being observed even at modest amount of alcohol.

The present finding of lower antibody titers among cigarette smokers agrees with those of previous studies^5-7^ and is also consistent with the mechanism for smoking-mediated immunopathology. Cigarette smoking induces chronic inflammation and downregulates CD4+ T cells and B cells, which are responsible for producing antigen-specific antibodies.^15^ Analyses of immunoglobulins confirmed a decreased production of IgA, IgG, and IgM in cigarettes smokers.^2^

To our knowledge, the present study is the first to show post-vaccination antibody titers in relation to HNB tobacco. We observed a decrease of antibody titers among users of HNB tobacco products relative to never smokers, although the reduction was less than that associated with cigarette smoking. This could be ascribed to the nicotine contained in HNB tobacco products, the amount of which in the fillers and the mainstream smoke is comparable to those of conventional cigarettes.^9, 16^ Experimental studies showed that chronic exposure to nicotine inhibited antibody-forming cell response, impaired antigen-mediated signaling in T cells, and induced T cell anergy.^15^ The present finding, together with experimental data, adds evidence to support the adverse effect of HNB tobacco product use against immunogenicity to the COVID-19 vaccination.

We found lower titers of anti-SARS-CoV-2 spike IgG antibodies among weekly drinkers of alcoholic beverages, a finding consistent with previous studies.^4^ The novelty of the present study is to show a clear dose-response relationship; antibody titers steadily decreased with increasing alcohol consumption, with significant reduction being observed even at moderate amount of alcohol. This finding is at variance with the hypothesis derived from mechanistic inference; moderate alcohol drinking can boost antibody response by decreasing the level of pathological inflammation and increasing circulating immunoglobulins and plasma antioxidants levels^10^. However, our observation is compatible with animal experiments showing that low doses of alcohol inhibited antigen-stimulated B-cells proliferation and their antibody production.^17, 18^ The present finding, together with experimental data, indicates that the detrimental effects of alcohol intake on the COVID-19 immunogenicity may start at the low doses without threshold.

Strengths of the present multi-center study include a large sample size, quantitative assessment of alcohol intake, information on the type of tobacco products, and the adjustment of multiple potential confounders. We should also acknowledge study limitations. First, as this study is cross-sectional, we cannot determine whether smoking and alcohol use were associated with the surge of antibodies in response to vaccination or the decay of antibodies over time. Second, the number of exclusive HNB tobacco users was not large enough to detect the reduction of IgG titers among them with statistical significance. Third, we did not measure neutralizing antibody, a more reliable marker of immunogenicity. Nevertheless, spike IgG antibody titers measured with the assay we employed are known to well correlate with neutralizing antibody titers (spearman’s ρ=0.71, 95% CI: 0.63–0.77).^19^ Finally, many East Asians are genetically deficient in one of the enzymes involved in metabolizing alcohol (aldehyde dehydrogenase);^11^ thus, the present finding regarding alcohol use may not be observed in populations with different genetic background.

In conclusion, cigarettes smokers and, to a lesser extent, HNB tobacco product users had lower IgG antibody titers against SAR-CoV-2 spike protein after vaccination, relative to never-smokers. The antibody titers steadily decreased with increasing alcohol consumption, with significant reduction being observed even at moderate amount of alcohol. In addition to conventional cigarette smoking and heavy alcohol use, the use of HNB tobacco products and moderate alcohol drinking may deteriorate immune response to COVID-19 vaccine.

## Ethics approval

Written informed consent was obtained from each participant. After completing the opt-out process, the survey data were anonymized and submitted to the study committee for pooled analysis. The study design and procedure for data collection were approved by the ethical committee of each center, while those of pooling study were approved by that of the National Center for Global Health and Medicine (NCGM) (the approved number: NCGM-G-004233).

## Data Availability

All data produced in the present study are available upon reasonable request to the authors.

## Author contributions

Conceptualization: TM; Methodology: KY, MKojima, MI, RO, KN, TM; Software: KM; Formal analysis: SY; Investigation: SY, AT, KI, NM, AN, HT, ST, JU, KY, MKojima, MI, RO, KN, MKonishi, TM; Resources: AT, KM; Data Curation: SY, MKonishi; Writing-Original Draft: SY, TM; Writing-Review & Editing: All authors; Visualization: SY; Supervision: NO, KY, MKojima, MI, RO, KN, TM; Project administration: TM; Funding acquisition: TM.

## Data availability

The data underlying this article cannot be shared publicly due to ethical restrictions and participant confidentiality concerns, but de-identified data are available from Dr. Mizoue (Department of Epidemiology and Prevention, Center for Clinical Sciences, National Center for Global Health and Medicine, Tokyo, Japan) to qualified researchers on reasonable request.

## Supplemental Appendix

**Supplemental Table 1.** Details of each national center’s survey

| National Center | Timing of the vaccination programs | Timing of the serological survey | No. of invited staff to the serological survey | No. of staff answered the questionnaire, (%) | No. of staff donated blood sampling, (%) | No. of participants, (%)<br>(No. of staff had data on either the questionnaire or blood sample) |
| --- | --- | --- | --- | --- | --- | --- |
| NCC | April to June 2021 | June to July 2021 | 2,432 | 613 (25%) | 607 (25%) | 619 (25%) |
| NCCHD | March to April 2021 | July 2021 | 1,808 | 1520 (84%) | 1517 (84%) | 1520 (84%) |
| NCGG | May to June 2021 | June 2021 | 878 | 711 (81%) | 800 (91%) | 800 (91%) |
| NCGM | March to May 2021 | June 2021 | 3,072 | 2699 (88%) | 2763 (90%) | 2,779 (90%) |
| Total |  |  | 8,190 | 5543 (68%) | 5687 (69%) | 5,718 (70%) |

NCC: National Cancer Center, Tokyo, Japan
NCCHD: National Center for Child Health and Development, Tokyo, Japan
NCGG: National Center for Geriatrics and Gerontology, Aichi, Japan
NCGM: National Center for Global Health and Medicine, Tokyo, Japan

**Supplemental Figure 1.**
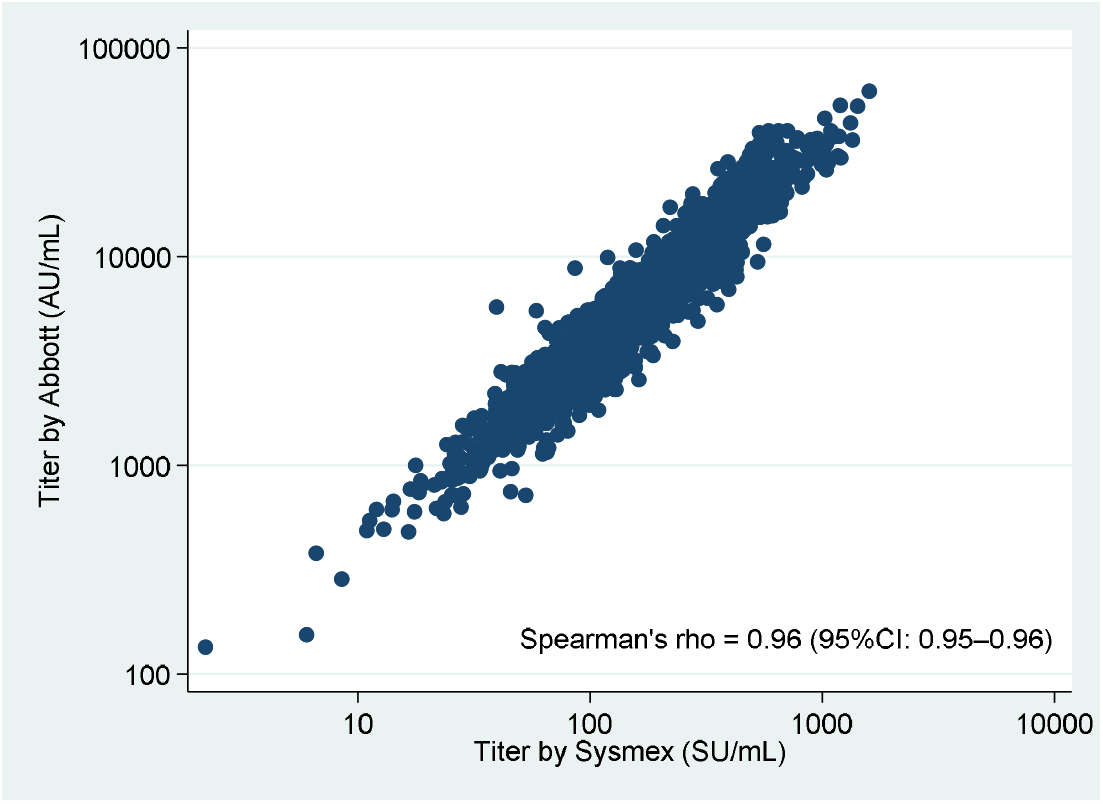
Scatter plot between anti-SARS-CoV-2 spike IgG antibody titers measured by Sysmex assay and Abbott assay among 2,961 healthcare workers.

**Supplemental Table 2.**
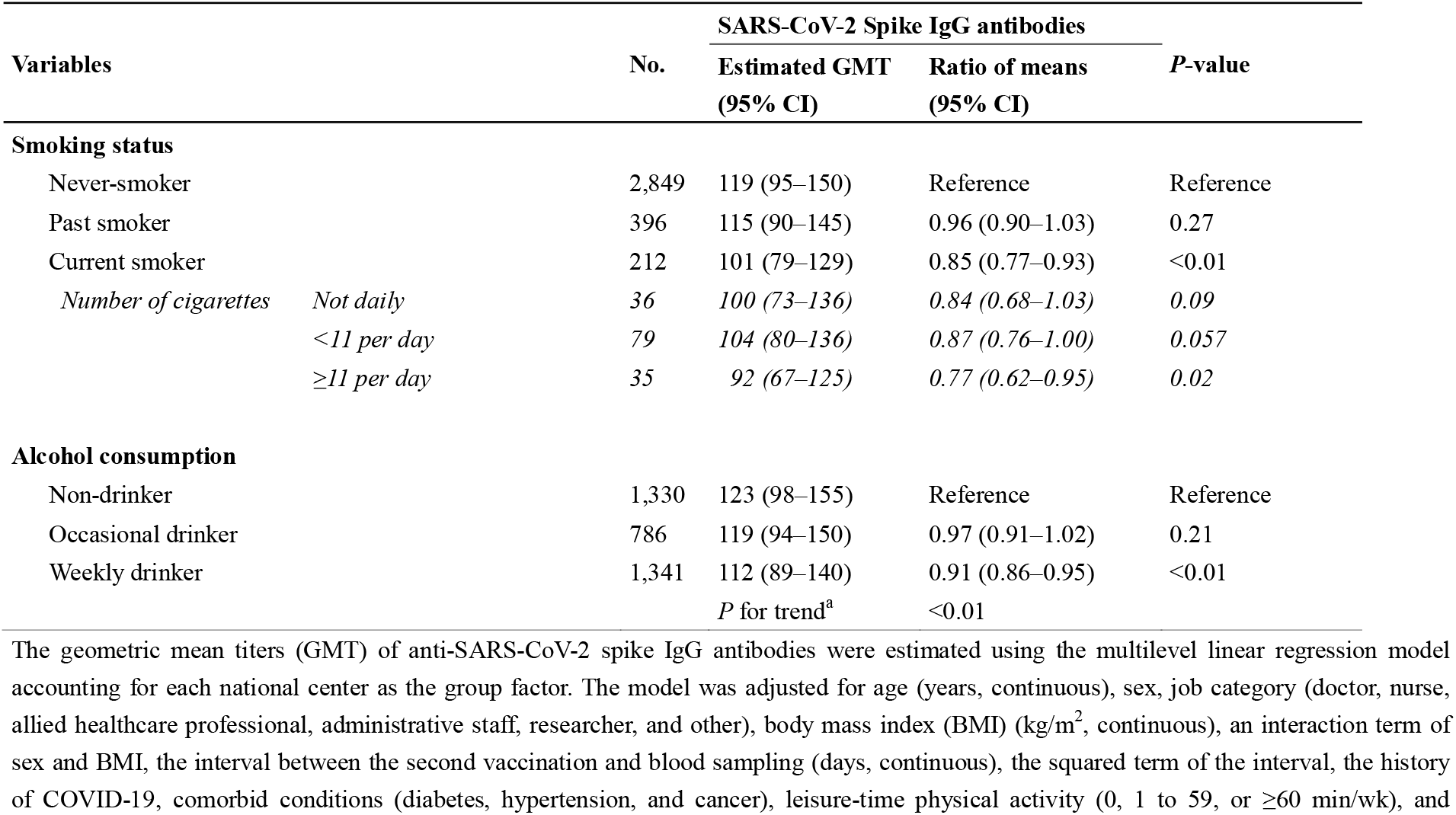
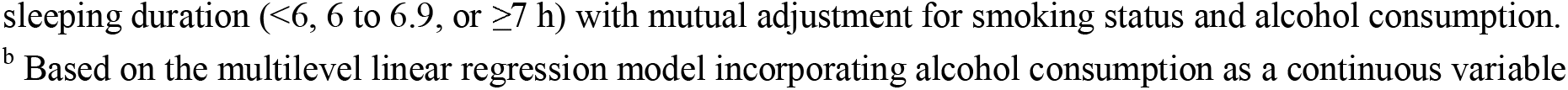
The estimated geometric means (GMT) with 95% confidence intervals (CI) according to smoking status (never-smoker, past smoker, and current smoker) and alcohol consumption (non-drinker, occasional drinker, and regular drinker) among 3,457 vaccinated healthcare workers.

| Variables |  | No. | SARS-CoV-2 Spike IgG antibodies |  | P-value |
| --- | --- | --- | --- | --- | --- |
|  |  |  | Estimated GMT<br>(95% CI) | Ratio of means<br>(95% CI) |  |
| Smoking status |  |  |  |  |  |
| Never-smoker |  | 2,849 | 119 (95–150) | Reference | Reference |
| Past smoker |  | 396 | 115 (90–145) | 0.96 (0.90–1.03) | 0.27 |
| Current smoker |  | 212 | 101 (79–129) | 0.85 (0.77–0.93) | <0.01 |
| Number of cigarettes | Not daily | 36 | 100 (73–136) | 0.84 (0.68–1.03) | 0.09 |
|  | <11 per day | 79 | 104 (80–136) | 0.87 (0.76–1.00) | 0.057 |
|  | ≥11 per day | 35 | 92 (67–125) | 0.77 (0.62–0.95) | 0.02 |
| Alcohol consumption |  |  |  |  |  |
| Non-drinker |  | 1,330 | 123 (98–155) | Reference | Reference |
| Occasional drinker |  | 786 | 119 (94–150) | 0.97 (0.91–1.02) | 0.21 |
| Weekly drinker |  | 1,341 | 112 (89–140) | 0.91 (0.86–0.95) | <0.01 |
|  |  |  | P for trend<sup>a</sup> | <0.01 |  |

The geometric mean titers (GMT) of anti-SARS-CoV-2 spike IgG antibodies were estimated using the multilevel linear regression model accounting for each national center as the group factor. The model was adjusted for age (years, continuous), sex, job category (doctor, nurse, allied healthcare professional, administrative staff, researcher, and other), body mass index (BMI) (kg/m<sup>2</sup>, continuous), an interaction term of sex and BMI, the interval between the second vaccination and blood sampling (days, continuous), the squared term of the interval, the history of COVID-19, comorbid conditions (diabetes, hypertension, and cancer), leisure-time physical activity (0, 1 to 59, or ≥60 min/wk), and
sleeping duration ( $<6$ , 6 to 6.9, or $\geq 7$ h) with mutual adjustment for smoking status and alcohol consumption.
<sup>b</sup> Based on the multilevel linear regression model incorporating alcohol consumption as a continuous variable

## References

1. Levin EG, Lustig Y, Cohen C, et al. Waning Immune Humoral Response to BNT162b2 Covid-19 Vaccine over 6 Months. N Engl J Med 2021.

2. Qiu F, Liang C-L, Liu H, et al. Impacts of cigarette smoking on immune responsiveness: Up and down or upside down? Oncotarget 2017; 8: 268–84.

3. Pasala S, Barr T, Messaoudi I. Impact of Alcohol Abuse on the Adaptive Immune System. Alcohol Res 2015; 37: 185–97.

4. Kageyama T, Ikeda K, Tanaka S, et al. Antibody responses to BNT162b2 mRNA COVID-19 vaccine and their predictors among healthcare workers in a tertiary referral hospital in Japan. Clin Microbiol Infect 2021.

5. Michos A, Tatsi E-B, Filippatos F, et al. Association of total and neutralizing SARS-CoV-2 spike -receptor binding domain antibodies with epidemiological and clinical characteristics after immunization with the 1st and 2nd doses of the BNT162b2 vaccine. Vaccine 2021; 39: 5963–7.

6. Nomura Y, Sawahata M, Nakamura Y, et al. Age and Smoking Predict Antibody Titres at 3 Months after the Second Dose of the BNT162b2 COVID-19 Vaccine. Vaccines 2021; 9: 1042.

7. Watanabe M, Balena A, Tuccinardi D, et al. Central obesity, smoking habit, and hypertension are associated with lower antibody titres in response to COVID-19 mRNA vaccine. Diabetes Metab Res Rev 2021.

8. Caputi TL, Leas E, Dredze M, Cohen JE, Ayers JW. They’re heating up: Internet search query trends reveal significant public interest in heat-not-burn tobacco products. PLoS One 2017; 12: e0185735.

9. Bekki K, Inaba Y, Uchiyama S, Kunugita N. Comparison of Chemicals in Mainstream Smoke in Heat-not-burn Tobacco and Combustion Cigarettes. Journal of UOEH 2017; 39: 201–7.

10. Messaoudi I, Pasala S, Grant K. Could moderate alcohol intake be recommended to improve vaccine responses? Expert Review of Vaccines 2014; 13: 817–9.

11. Wall TL, Ehlers CL. Genetic Influences Affecting Alcohol Use Among Asians. Alcohol Health Res World 1995; 19: 184–9.

12. Noda K, Matsuda K, Yagishita S, et al. A novel highly quantitative and reproducible assay for the detection of anti-SARS-CoV-2 IgG and IgM antibodies. Sci Rep 2021; 11.

13. Yamamoto S, Mizoue T, Tanaka A, et al. Sex–associated differences between body mass index and SARS-CoV-2 antibody titers following the BNT162b2 vaccine among 2,435 healthcare workers in Japan. medRxiv (preprint) 2021.

14. Desquilbet L, Mariotti F. Dose-response analyses using restricted cubic spline functions in public health research. Stat Med 2010: 1037–57.

15. Sopori M. Effects of cigarette smoke on the immune system. Nature Reviews Immunology 2002; 2: 372–7.

16. Simonavicius E, McNeill A, Shahab L, Brose LS. Heat-not-burn tobacco products: a systematic literature review. Tob Control 2019; 28: 582–94.

17. Aldo-Benson M. Mechanisms of alcohol-induced suppression of B-cell response. Alcohol Clin Exp Res 1989; 13: 469–75.

18. Aldo-Benson M, Kluve-Beckerman B, Hardwick J, Lockwood M. Ethanol inhibits production of messenger ribonucleic acid for kappa-chain in stimulated B lymphocytes. J Lab Clin Med 1992; 119: 32–7.

19. Maeda K, Amano M, Uemura Y, et al. Correlates of Neutralizing/SARS-CoV-2-S1-binding Antibody Response with Adverse Effects and Immune Kinetics in BNT162b2-Vaccinated Individuals. medRxiv (preprint) 2021.

